# COVID-19 Vulnerability of Transgender Women With and Without HIV Infection in the Eastern and Southern U.S.

**DOI:** 10.1101/2020.07.21.20159327

**Authors:** Tonia C. Poteat, Sari L. Reisner, Marissa Miller, Andrea L. Wirtz, on behalf of the American Cohort To Study HIV Acquisition Among Transgender Women (LITE)

## Abstract

**Background:** COVID-19 is a new global pandemic and people with HIV may be particularly vulnerable. Gender identity is not reported, therefore data are absent on the impact of COVID-19 on transgender people, including transgender people with HIV. Baseline data from the American Cohort to Study HIV Acquisition Among Transgender Women in High Risk Areas (LITE) Study provide an opportunity to examine pre-COVID vulnerability among transgender women.

**Setting:** Atlanta, Baltimore, Boston, Miami, New York City, Washington, DC

**Methods:** Baseline data from LITE were analysed for demographic, psychosocial, and material factors that may affect risk for COVID-related harms.

**Results:** The 1020 participants had high rates of poverty, unemployment, food insecurity, homelessness, and sex work. Transgender women with HIV (n=273) were older, more likely to be Black, had lower educational attainment, and were more likely to experience material hardship. Mental and behavioural health symptoms were common and did not differ by HIV status. Barriers to healthcare included being mistreated mistreatment, uncomfortable providers, and past negative experiences; as well as material hardships, such as cost and transportation. However, most reported access to material and social support – demonstrating resilience.

**Conclusions:** Transgender women with HIV may be particularly vulnerable to pandemic harms. Mitigating this harm would have positive effects for everyone, given the highly infectious nature of this coronavirus. Collecting gender identity in COVID-19 data is crucial to inform an effective public health response. Transgender-led organizations’ response to this crisis serve as an important model for effective community-led interventions.

## Introduction

In a few short months, more than 2.5 million people in the United States (US) have tested positive for SARS-CoV-2, the novel coronavirus responsible for COVID-19; and more than 128,00 people have died.^1^ The impact of COVID-19 on people with HIV is not yet known. However, individuals with immune compromise are more vulnerable to severe illness.^2^ Marked racial and ethnic disparities exist for both COVID-19 and HIV. Black communities bear the brunt of these diseases as well as the underlying conditions associated with negative COVID-19 outcomes.^2,3^ Latinx populations have some of the highest SARS-CoV-2 positivity rates (43%)^4^ and represent one-fourth of new HIV diagnoses in the US, despite representing 17% of the US population.^5^

Social conditions play an important role in COVID-19 vulnerability. Many essential workers earn low wages and face increased risk for SARS-CoV-2 exposure.^6^ Sex workers in the underground economy cannot maintain social distancing and also work to meet their economic needs.

While data on race are now reported for COVID-19,^7^ only one state plans to collect data disaggregated by gender identity.^8^ The American Cohort to Study HIV Acquisition Among Transgender Women in High Risk Areas (LITE) Study provides an opportunity to examine pre-COVID conditions that may impact transgender women’s socioeconomic, health, and mental health outcomes following this crisis.^9^

LITE is a longitudinal study of transgender women at risk for HIV. Facility-based participants were recruited via convenience sampling in Atlanta, Baltimore, Boston, Miami, New York City, and Washington, DC. Eligibility included identity as a woman or along a transfeminine spectrum, male sex assignment at birth, and age 18 years and older. Participants self-administered a survey about demographics, mental and behavioural health, material hardship, sex work, and social support; and they provided biological samples for HIV testing. We analysed data from baseline study visits conducted from study onset in March 2018 to March 2020 when WHO declared COVID-19 a pandemic. We compared pre-pandemic experiences by baseline HIV status.

A total of 1,020 transgender women completed the baseline assessment, among whom 27% had HIV. Participants with HIV were older, more likely to be Black, and had lower educational attainment. There were no differences in ethnicity or immigration by HIV status.

### Material Hardship

COVID-19 and efforts to contain it may induce serious harm among socioeconomically vulnerable people, including transgender women.^10,11^ Overall, more than half of LITE participants were unemployed (53.6%, n=547); 46% had incomes below the federal poverty level (n=470); 13% had been homeless in the prior 3 months (n=131); and 21% (n=207) had engaged in sex work in the prior 3 months. Almost half of LITE participants were food insecure (48.2%; n=488), almost 5-fold that of the general US population.^12^ Material hardship were even greater for transgender women with HIV, who were significantly more likely than those without HIV to be unemployed (76.6% vs. 45.2%; p<0.001), have public insurance (88.6% vs. 48.5% (p<0.001), earn an income below the federal poverty level (66.3% vs. 38.7%, p<0.001), engage in sex work (30.3% vs. 17.2%; p<0.001), experience food insecurity (63.7% vs. 42.5%; p<0.001), and be homeless in the prior 3 months (17.5% vs. 11.4%; p=0.02).

High baseline rates of unemployment added to COVID-19-related job losses may push transgender women even deeper into poverty, exacerbating food insecurity and likely increasing reliance on sex work. Sex work may then increase transgender women’s risk for acquiring COVID-19. Transgender women who were already engaged in sex work may have a reduced income due to social distancing. However, criminalization of sex work precludes access to economic relief from federal funds while also increasing the risk of incarceration where again, they face elevated risk of COVID-19.^13^

The high rate of homelessness among transgender women also puts them at substantial risk for COVID-19. Regular handwashing and social distancing may be impossible without a home. Transgender women seeking refuge in the sex-segregated shelter system often face discrimination and outright denial of services. If they are able to access shelter services, they may face crowded conditions that increase COVID-19 risk. Given national shortages of personal protective equipment for essential workers, it is unlikely that transgender women in shelters will have means to protect themselves from COVID-19.

### Psychosocial vulnerabilities

Overall, transgender women in LITE reported high pre-COVID levels of psychosocial vulnerability. Psychological distress was common, with more than a quarter of participants scoring 13 or higher on the Kessler 6 (27.4%; n=279). A remarkable 41% reported symptoms indicative of post-traumatic stress disorder (PTSD; n=417) and 28% reported suicidal ideation in the prior 6 months (n=279). Transgender women with HIV were less likely to report these symptoms than HIV-negative transgender women (**Figure 1**). This difference may be explained by access to mental health services available to people with HIV through federal Ryan White HIV Care Act funding. More than one-quarter of participants reported symptoms of alcohol (29.0; n=296) and substance use disorder (29.4%; n=294), with no difference by HIV status. The overall prevalence of mental health symptoms in LITE exceed the 19% prevalence of any mental illness found in the general population.^14^ The prevalence of PTSD symptoms is twice that reported in primary care samples (23%) and rivals rates found among Vietnam War veterans (31%).^15,16^

**Figure 1.**
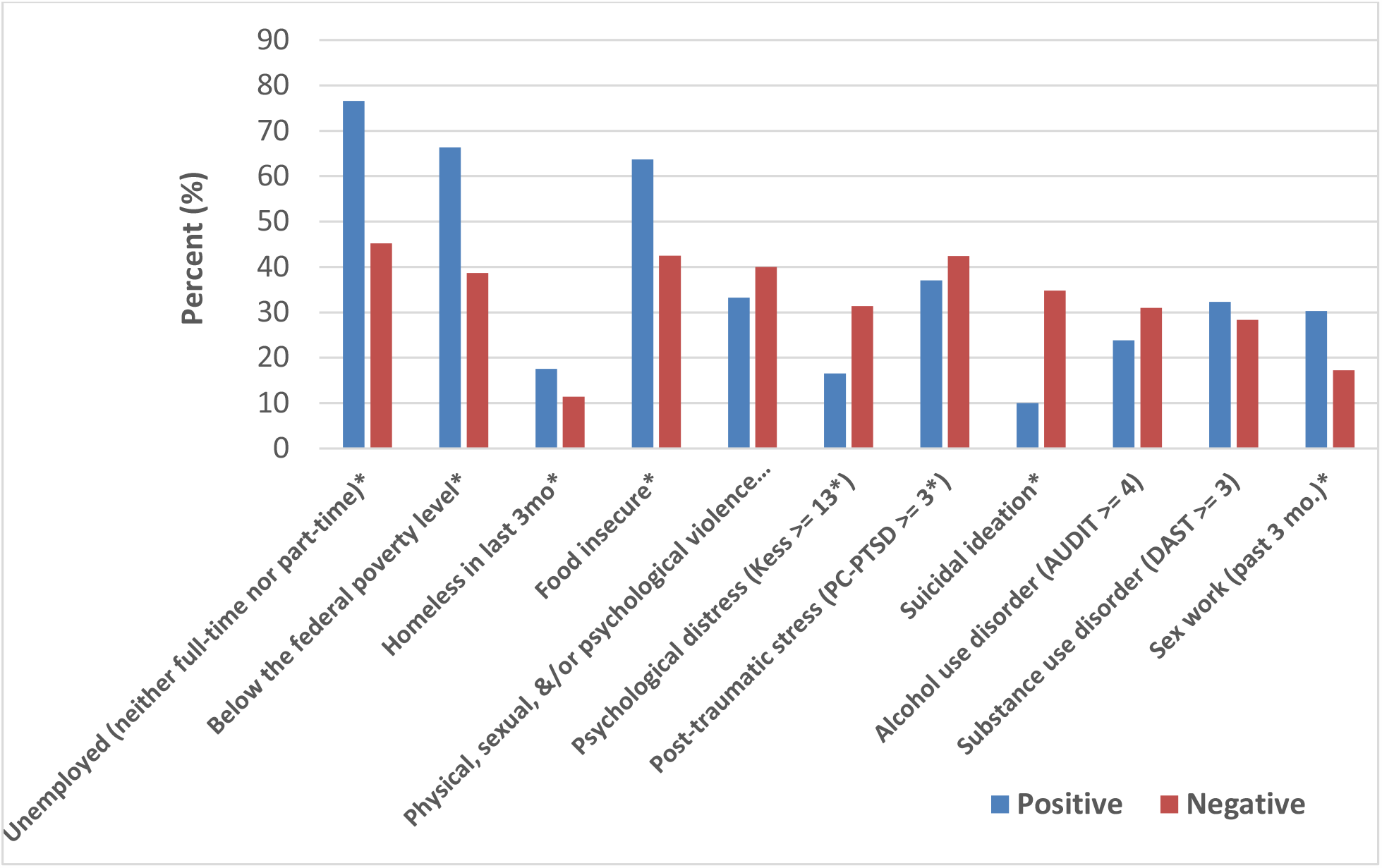
Pre-COVID-19 Socioeconomic and Psychosocial Characteristics by Laboratory-Confirmed HIV Status at Baseline in the LITE Study

In response to the COVID-19 crisis, state and local governments have encouraged social distancing, closed non-essential businesses and schools, prohibited large gatherings, and declared mandatory stay-at-home orders for all but essential workers. Social distancing measures, while critical to curbing the epidemic, have isolated many people with alcohol and substance use disorders (AUD and SUD) from treatment programs and 12-step groups, increasing the risk for relapse. In addition, some individuals may use alcohol and drugs to cope with coronavirus stress or stress of isolation. Prior to the pandemic, transgender women in this study reported almost five times the national rate of AUD (5.8%) and three times the rate of SUD (8.9%), suggesting transgender women may be vulnerable to worsening behavioral health from COVID-19 stress.^17^

Recent data indicate that a significantly higher proportion of people who were sheltering in place (47%) reported negative mental health effects of coronavirus stress than people who were not sheltering in place (37%).^18^ Negative mental health effects due to social isolation and stress may be particularly pronounced for transgender women with HIV who are already at high risk for distress and suicidal ideation. Similar stressors may exist for perpetrators of violence. Before COVID-19, 38.2% of participants reported some form of gender-based violence within the prior 3 months, and these levels may climb as transgender women are forced to isolate with abusive partners or others.

### COVID-19 and access to healthcare

Despite lack of available data on COVID-19 by gender identity, social and structural vulnerabilities suggest transgender women, particularly transgender women with HIV, may be disproportionately impacted by the pandemic. Lack of employment, reliance on public insurance, and higher probability of living in an urban area mean that transgender women who develop COVID-19 symptoms will have few choices for care beyond crowded urban hospitals which may already be overwhelmed with COVID-19 patients.

LITE participants reported frequent barriers to healthcare, including mistreatment for being transgender (19.3%), a provider who was uncomfortable caring for transgender patients (28.9%), bad experiences in the past (35.7%), as well as significant challenges related to material hardship, such as cost (43%) and transportation (43.7%). Several reports have documented reductions in emergency care visits due to concerns of acquiring COVID-19 in health facilities;^19,20^ such concerns coupled with unique barriers to care may lead transgender women to delay care-seeking for COVID-19, HIV, or other conditions until symptoms are severe, thus increasing their risk for negative outcomes or death.

### Support and community resilience

In the face of significant adversities, transgender women exhibit remarkable resilience. In LITE, most transgender women could identify someone who provided them with emotional and material support. Sixty-four percent (n=652) had someone to care for them if they were sick, and 60% (n=611) had someone who could lend them money. This support was present regardless of HIV status. Such mutual support has come to the fore in the wake of the COVID-19 pandemic, as numerous transgender-led organizations have stepped up to provide both social and material support, including rapid response funding for community members in need.^21-23^

## Conclusions

Transgender women with HIV may be particularly vulnerable to harms associated with COVID-19 due to precarious access to employment, income, food, housing, and heightened vulnerability to violence. Evolving national policies have the potential to reduce or increase vulnerability. In June 2020, the US Supreme Court ruled that Title VII of the Civil Rights Act of 1964 protects transgender people against employment discrimination,^24^ creating opportunities to reduce socioeconomic vulnerabilities. Yet, a week prior, the Department of Health and Human Services excluded gender identity from protections against sex discrimination in healthcare,^25^ creating another barrier to care. Given the highly contagiousness nature of SARS-CoV-2, understanding and mitigating its impact on vulnerable communities will benefit everyone. Collecting gender identity in COVID-19 surveillance data will be crucial to inform public health responses. Transgender-led organizations’ response to this crisis serve as an important model for effective community-led interventions.

## Data Availability

Data are available from the LITE Study upon request. Requests for data must be accompanied by a concept note submitted via https://www.litestudy.org and must be consistent with the LITE Study collaboration policies listed on the website.

## Authors’ contributions

TP conceived of the manuscript and led writing. AW and SR led data collection activities. AW conducted statistical analyses and MM reviewed results for face validity. All authors contributed to reviewing and editing drafts of the manuscript.

## Author information

Among co-authors, diversity existed along lines of gender identity, race, and sexual orientation. Two authors identify as transgender, two identify as Black, and two identify as queer. All authors are deeply committed to the health and well-being of transgender communities and strive to conduct research that advances health equity.

## Acknowledgements

The authors would like to express their gratitude to the transgender women who take part in this study. This study would not be possible without their participation. We also acknowledge the work of the entire American Cohort To Study HIV Acquisition Among Transgender Women team: Andrea Wirtz (multiple PI; JHU); Sari Reisner (multiple PI; Harvard University); Keri Althoff (JHU); Chris Beyrer (JHU); James Case (JHU); Erin Cooney (JHU); Oliver Laeyendecker (JHU); Kathleen Powers (JHU) and Jeffrey Herman (JHU); Tonia Poteat (University of North Carolina); Kenneth Mayer (Fenway Health); Asa Radix (Callen-Lorde Community Health Center); Christopher Cannon (Whitman-Walker Health); W. David Hardy (Whitman-Walker Health); Jason Schneider (Emory University and Grady Hospital); Sonya Haw (Emory University and Grady Hospital); Allan Rodriguez (University of Miami); Andrew Wawrzyniak (University of Miami); the incredible research teams at each study site; and the LITE community advisory board, including the following individuals: Jennifer Lopez, Sherri Meeks, Sydney Shackelford, Nala Toussaint, SaVanna Wanzer, and Joseph Zolobczuk, as well as those who have remained anonymous.

## References

1. Johns Hopkins University. COVID-19 Dashboard. https://coronavirus.jhu.edu/map.html. Accessed.

2. Centers for Disease Control and Prevention. Cases of Coronavirus Disease (COVID-19) in the U.S. https://www.cdc.gov/coronavirus/2019-ncov/cases-updates/cases-in-us.html. Accessed April 24, 2020.

3. Marron MM, Ives DG, Boudreau RM, Harris TB, Newman AB. Racial Differences in Cause-Specific Mortality Between Community-Dwelling Older Black and White Adults. J Am Geriatr Soc. 2018;66(10):1980–1986.

4. Martinez DA, Hinson JS, Klein EY, et al. SARS-CoV-2 Positivity Rate for Latinos in the Baltimore-Washington, DC Region. JAMA. 2020.

5. Centers for Disease Control and Prevention. Estimated HIV incidence and prevalence in the United States, 2010–2015. HIV Surveillance Supplemental Report. 2018.

6. Jin B, McGill A. Who is most at risk in the coronavirus crisis: 24 million of the lowest-income workers. Politico. https://www.politico.com/interactives/2020/coronavirus-impact-on-low-income-jobs-by-occupation-chart/. Updated March 22, 2020. Accessed April 26, 2020.

7. Millett GA, Jones AT, Benkeser D, et al. Assessing Differential Impacts of COVID-19 on Black Communities. Annals of epidemiology. 2020.

8. Gov. Wolf Announces Inclusion of Gender Identity, Sexual Orientation or Expression in COVID-19 Data Collection. Commonwealth of Pennyslvania. https://www.governor.pa.gov/newsroom/gov-wolf-announces-inclusion-of-gender-identity-sexual-orientation-or-expression-in-covid-19-data-collection/. Updated May 13, 2020. Accessed June 29, 2020.

9. Wirtz AL, Poteat T, Radix A, et al. American Cohort to Study HIV Acquisition Among Transgender Women in High-Risk Areas (The LITE Study): Protocol for a Multisite Prospective Cohort Study in the Eastern and Southern United States. JMIR Res Protoc. 2019;8(10):e14704.

10. United Nations DoEaSA. The Social Impact of COVID-19. https://www.un.org/development/desa/dspd/2020/04/social-impact-of-covid-19/. Published 2020. Updated April 6, 2020. Accessed April 26, 2020.

11. Herman JL, O’Neill K. Vulnerabilities to COVID-19 Among Transgender Adults in the U.S. Williams Institute. https://www.un.org/development/desa/dspd/2020/04/social-impact-of-covid-19/. Published 2020. Updated April 2020. Accessed April 26, 2020.

12. USDA. Food Security Status of U.S. Households in 2018. https://www.ers.usda.gov/topics/food-nutrition-assistance/food-security-in-the-us/key-statistics-graphics.aspx. Published 2019. Accessed June, 2020.

13. Hurley M. Why prisoners are at higher risk for the coronaviru: 5 questions answered. The Conversation. https://theconversation.com/why-prisoners-are-at-higher-risk-for-the-coronavirus-5-questions-answered-136111. Published 2020. Updated April 17, 2020. Accessed April 26, 2020.

14. Results from the 2017 National Survey on Drug Use and Health. Substance Abuse and Mental Health Services Administration. https://www.samhsa.gov/data/sites/default/files/cbhsq-reports/NSDUHDetailedTabs2017/NSDUHDetailedTabs2017.htm#tab8-33A. Published 2017. Accessed April 26, 2020.

15. Liebschutz J, Saitz R, Brower V, et al. PTSD in urban primary care: high prevalence and low physician recognition. J Gen Intern Med. 2007;22(6):719–726.

16. Gradus JL. Epidemiology of PTSD. U.S. Department of Veterans Affairs, PTSD: National Center for PTSD. https://www.ptsd.va.gov/professional/treat/essentials/epidemiology.asp#two. Accessed April 26, 2020.

17. Substance Abuse and Mental Health Services Administration. Natoinal Survey on Drug Use and Health. https://www.samhsa.gov/data/sites/default/files/cbhsq-reports/NSDUHDetailedTabs2018R2/NSDUHDetTabsSect5pe2018.htm#tab5-4b. Published 2018. Accessed April 26, 2020.

18. Kirzinger A, Kearney A, Hamel L, Brodie M. KFF Health Tracking Poll - Early April 2020: The Impact of Coronavirus on Life In America. In: Kaiser Family Foundation; 2020.

19. McFarling UL. “Where are all our patients?’: Covid phobia is keeping people with serious heart symptoms away from ERs. STAT News. https://www.statnews.com/2020/04/23/coronavirus-phobia-keeping-heart-patients-away-from-er/?utm_source=STAT+Newsletters&utm_campaign=acc8d5e249-Weekend_Reads_COPY_02&utm_medium=email&utm_term=0_8cab1d7961-acc8d5e249-149836233. Published 2020. Updated April 23, 2020. Accessed April 26, 2020.

20. Garcia S, Albaghdadi MS, Meraj PM, et al. Reduction in ST-Segment Elevation Cardiac Catheterization Laboratory Activations in the United States during COVID-19 Pandemic. Journal of the American College of Cardiology. 2020.

21. Black Trans COVID-19 Community Response https://blacktrans.org/covid-19-volunteers/. Accessed April 26, 2020.

22. COVID-19 Trans Resources Directory. https://translash.org/covid-19-trans-resources-directory/. Accessed April 26, 2020.

23. Solutions T. COVID-19 Relief Rapid Response. https://www.transsolutionsconsulting.org/covid-19-relief?fbclid=IwAR3upkLpagxu9Gj_xehLIJZ2i882AEzyFFmjfx4lkNZzDjymujm87ztod64. Accessed April 26, 2020.

24. Robb M. Supreme Court Ruling Protects LGBTQ and Transgender Employees against Discrimination. The National Law Review. 2020;X.

25. Simons-Duffin S. Transgender Health Protections Reversed By Trump Administration. NPR,. 12 June 2020, 2020.

